# Breath Acetone Correlates with Capillary β-hydroxybutyrate in Type 1 Diabetes

**DOI:** 10.1101/2025.01.30.25321320

**Authors:** Kai E. Jones, Max C. Petersen, Alexander M. Markov, Maamoun Salam, Petra Krutilova, Alexis M. McKee, Kathryn L. Bohnert, Samantha E. Adamson, Janet B. McGill

## Abstract

**Background:** Breath acetone (BrACE) is an end product of ketone metabolism that is measurable by noninvasive breath ketone analyzers. We assessed the correlation between capillary blood β-hydroxybutyrate (BOHB) and BrACE in people with type 1 diabetes (T1D) during 14 days of outpatient care with and without dapagliflozin treatment and during supervised insulin withdrawal studies with and without dapagliflozin.

**Methods:** In this randomized crossover study, participants completed 14-day two outpatient periods with or without dapagliflozin 10 mg daily. Each 14-day unsupervised outpatient period was followed by a one-day supervised insulin withdrawal study. Paired BOHB and BrACE measurements were obtained three times daily during outpatient periods, then hourly during supervised insulin withdrawal. The correlation between BrACE and BOHB was assessed by Spearman’s ρ.

**Results:** Twenty people with T1D completed the study. During outpatient periods, BrACE and BOHB were moderately correlated (n=1425 paired readings; ρ = 0.41; 95% CI: 0.36 to 0.45; *P* < 0.0001). However, BrACE and BOHB were strongly correlated during insulin withdrawal (n=246 paired values, ρ = 0.81; 95% CI: 0.77 to 0.85). In ROC analysis, BrACE > 5 ppm demonstrated optimal sensitivity (93%) and specificity (87%) for detecting capillary BOHB ≥ 1.5 mmol/L. No serious adverse events occurred.

**Conclusions:** In adults with T1D, measurement of breath acetone provides a noninvasive estimate of blood BOHB concentration. The correlation between BrACE and BOHB was suboptimal during unsupervised outpatient care, but was strong during supervised insulin withdrawal.

Trial registration: clinicaltrials.gov (NCT05541484)

## Introduction

Despite progress in developing insulin analogs and insulin delivery devices, the frequency of hospitalizations due to diabetic ketoacidosis (DKA), the most critical acute complication of type 1 diabetes (T1D), has risen.^1^ Furthermore, the mortality risk associated with DKA has not improved over the past decade.^1–3^ Risk factors for DKA include poor adherence to insulin regimens, errors in insulin administration, chronic poor glucose control, acute illness and use of sodium glucose co-transporter 2 inhibitors (SGLT2i).^4,5^ Individuals with T1D are instructed to measure ketones in the setting of symptoms of ketosis, during other illnesses, and when severe hyperglycemia occurs.^4,6,7^ Although routine ketone monitoring during usual care is generally not recommended, this has been shown to identify individuals at higher risk of DKA.^8^ In practice, ketone monitoring remains suboptimal; over 50% of adults do not test for ketones during periods of illness.^9–11^

Sodium-glucose cotransporter 2 inhibitors (SGLT2i) improve glycemic control, decrease insulin requirements, and reduce glycated hemoglobin levels in T1D.^12–18^ However, multiple randomized trials and meta-analyses have revealed a greater incidence of DKA and higher ketone levels in individuals using SGLT2i.^13,14,19,20^ Ketone monitoring has been proposed as a strategy for DKA mitigation in the setting of SGLT2i use.^12,21–24^

Ketone monitoring refers to the routine surveillance of one of the three major circulating ketone bodies: acetoacetate, β-hydroxybutyrate (BOHB), or acetone.^25^ Acetoacetate is measured in the urine using semi-quantitative test strips, whereas BOHB can be measured quantitatively by point-of-care capillary blood ketone meters. Urine acetoacetate testing has major limitations: it measures the average ketone concentration in urine stored in the bladder since the previous void, and urinary ketone levels can misleadingly increase when treatment for ketosis is initiated as BOHB is oxidized to acetoacetate.^26^ Capillary blood ketone meters that measure BOHB are commercially available but the technology is not widely used due to expense, inconvenience, and possibly inexperience with ketone meters.^24^

Measurement of breath acetone (BrACE), the highly volatile metabolic end product of ketone metabolism excreted through the lung, has not been thoroughly evaluated in persons in T1D. Breath ketone analyzers (BKA) have been employed in non-diabetic individuals to monitor adherence to ketogenic diets. Previous studies have demonstrated linear relationships between BrACE and plasma acetone during episodes of DKA ^27^, and between BrACE and blood BOHB in various settings, suggesting potential utility of BrACE in risk prediction for ketosis and DKA.^28–30^ Breath acetone meters are subject to potential interference from volatile organic compounds (VOCs) including alcohols, acetic acid, propionic acid, and butyric acid. Commonly used household items like cleaning products, nail polish removers, hand sanitizer, lip balm and fermented foods contain VOCs which can lead to falsely elevated readings of breath acetone. Recent food or alcohol ingestion or smoking can also falsely elevate BrACE readings.

The Biosense breath ketone analyzer (Readout Health) has previously been studied in people without diabetes who were instructed to follow either a standard diet or a ketogenic diet and collected paired capillary BOHB and BrACE measurements five times daily. A moderate correlation (r = 0.75) between BrACE and BOHB was observed in this setting.^31^ In a study investigating the use of a breath acetone analyzer in adults and children with T1D, a significant correlation between BrACE and capillary blood ketones was observed in adults but not in children.^32^ BrACE >3.9 ppm yielded optimal performance for detecting BOHB >0.6 mmol/L, with sensitivity of 94.7% but specificity of just 54.2%.^32^ In this context, breath ketone analyzers are not currently recommended by professional society guidelines for ketone monitoring in those with T1D.

The goal of this study was to assess the utility of BKA to detect mild to moderate ketosis in persons with T1D and to compare BrACE levels with capillary BOHB. The correlation between capillary BOHB and BrACE was assessed in unsupervised outpatient settings with and without an SGLT2i and during insulin withdrawal to induce mild to moderate ketosis. To further evaluate the utility of BrACE in identifying a threshold level of ketosis that would require intervention, we interrogated the data using receiver-operating characteristic (ROC) analysis to identify the BrACE level that best predicted capillary blood BOHB concentration ≥1.5 mmol/L. This point-of-care capillary blood BOHB threshold has been validated to identify impending DKA, demonstrating a sensitivity and specificity ranging from 93-100% and 85-98%, respectively.^33–35^

## Methods

### Study Design

This was an open-label, single-center (Washington University School of Medicine, St. Louis, MO) randomized crossover trial in which study participants completed two study periods in random order: usual care (UC) and usual care plus dapagliflozin (DAPA). During UC, participants continued their usual insulin regimen, either multiple daily injections (MDI) or continuous subcutaneous insulin infusion (CSII), for 14 days while performing paired measurements of capillary BOHB and BrACE 3 times per day. This was followed by a one-day supervised insulin withdrawal study. During DAPA, participants added dapagliflozin 10 mg daily to their usual care for 14 days and continued to perform paired measurements of capillary BOHB and BrACE 3 times per day. A supervised insulin withdrawal study followed immediately after a dose of dapagliflozin in the morning. The study was unblinded to both participants and investigators.

### Study Participants

The study was approved by the Washington University Human Research Protection Office (IRB #202206078) and all study participants provided written informed consent. The trial was registered in clinicaltrials.gov (NCT05541484). Inclusion criteria included: i) T1D of >1 year duration, ii) age 18 to 75 years, iii) HbA1c ≤10%, iv) stable insulin delivery method for the past 30 days, v) vision of 20/40 or better, vi) insulin delivery by either continuous subcutaneous insulin infusion (CSII) or multiple daily injections of insulin (MDI), and vii) use of Dexcom G6 or G7 continuous glucose monitor (CGM). Key exclusion criteria include: i) history of DKA within 6 months of study entry or more than 1 episode of DKA in the past 2 years, ii) use of SGLT2i or prior intolerance of SGLT2i, iii) eGFR <30 mL/min/1.73 m^2^, iv) adherence to a very-low-carbohydrate (<90 g/day) or ketogenic diet, v) pregnancy, plan to become pregnant within the next 3 months, or lack of contraceptive use in premenopausal women, vi) history of urinary tract infection within 3 months of study entry, vii) unwilling to avoid alcohol ingestion during the study period, viii) vomiting within the past 30 days.

### Breath Ketone Analysis

The Biosense® breath ketone analyzer uses a metal oxide semiconductor (MOS) sensor which is selective for acetone. A specific breathing pattern is recommended that includes prolonged forced exhalation because breath acetone concentration increases during exhalation. The MOS sensor is housed inside a sealed flow cell to decrease the entrance of surrounding environmental air.^31^ Results from this device are reported as units of parts per million (ppm). Each device is calibrated using laboratory gas standards certified by the National Institute of Standards and Technology, with acetone concentrations of 0, 5, and 20 ppm.

### Procedures

Study participants completed a screening evaluation including demographics, medical history, physical examination, assessment of insulin administration (doses and routes), download of continuous glucose monitor (CGM) data and insulin pump data if applicable. Participants were instructed in the use of the Precision Xtra® ketone monitoring system and the Biosense® breath ketone analyzer (BKA), and the need for paired (within 30 minutes) measurements three times every day. Specifically, participants were instructed to use the BKA device prior to eating, smoking, drinking soda, using toothpaste, mouthwash, or lip balm and to avoid environmental contaminants such as cleaning solutions. The device was to be used only in indoor spaces only. Study participants were educated on a ketone action plan and provided individual insulin dosing recommendations for capillary blood BOHB ≥ 1.5 mmol/L. During the DAPA period, insulin dose adjustments were recommended based on insulin delivery method and level of glucose control. The standard recommendation was to reduce bolus doses by 10-20%.

For the insulin withdrawal study, participants were admitted to the Clinical Translational Research Unit (CTRU) after an overnight fast. Participants on continuous subcutaneous insulin infusions were instructed to stop insulin delivery 1 hour prior to arrival. Participants using multiple daily injections (MDI) were transitioned to morning basal insulin for the study period, which was held the morning of the insulin withdrawal. Participants were required to have blood glucose level between 90-250 mg/dL and capillary blood BOHB <1.5 mmol/L before leaving home on the morning of the supervised insulin withdrawal study visit or the visit was cancelled. Participants remained fasting with serial venous blood sampling for plasma BOHB and basic metabolic panel every 2 hours and paired measurements of capillary blood glucose, capillary blood BOHB, and BrACE every hour. A plastic cap was applied to the BKA device when not in use to minimize VOC interference. The supervised insulin withdrawal study was terminated when one or more stopping criteria were met (capillary blood glucose > 400 mg/dL, capillary blood BOHB > 4 mmol/L, symptoms of DKA, more than 8 hours elapsed from insulin withdrawal, or participant request).

### Outcomes

The primary outcome for the study was the correlation coefficient between capillary blood BOHB and BrACE in the outpatient periods with and without dapagliflozin. Other outcomes included the correlations between capillary blood BOHB and BrACE levels during the insulin withdrawal days. Adverse events were actively collected for safety and included the daily ketone measurements, severe hypoglycemia events, sensor glucose <54 mg/dL for more than 1% of time, diabetic ketoacidosis, genitourinary infections, gastrointestinal distress, or dehydration.

### Statistical Analysis

The prespecified primary outcome of the study was the correlation of capillary blood BOHB and BrACE during the 2-week outpatient periods. We calculated using the pwr R package that at least 84 ketone measurements above baseline (∼0.1 mmol/L or 1 BrACE ppm would provide >80% power to detect a weak correlation of 0.3 using a two-sided significance level of 0.05 and estimated that 20 participants would need to be enrolled to achieve this sample size.^31^ Descriptive statistics were examined for normality by using histograms and Kolmogorov-Smirnov tests and are reported as mean ± SD or median (interquartile range) as appropriate. Median capillary blood BOHB or BrACE concentrations between UC and DAPA periods were compared by using the Mann-Whitney U test. Correlations of blood and breath ketone concentrations were calculated as Spearman’s ρ given non-normal variable distributions. For ROC curves, Youden’s J statistic was used to determine the optimal BrACE threshold. Values were excluded from analysis if study participants described reason for BrACE >10 ppm that included food or drink prior to measurement, use of lip balm, environmental contaminants or measurements taken outside. Paired values were compared by using paired two-tailed t-tests or Wilcoxon matched-pairs signed-rank test as appropriate. Statistical analyses were completed in SPSS 29 (IBM), R version 4.3.2 and GraphPad Prism 10.

## Results

Participant characteristics are shown in **Table 1**. Twenty-one adults with T1D were screened, and 20 participated in the study, including 11 males and 9 females. The average age was 48 ± 18 years and mean BMI was 29.5 ± 5.2 kg/m^2^. CSII in hybrid closed loop systems were used by 16 (80%) of the participants. At baseline, CGM time-in-range (70-180 mg/dL) was 61 ± 18%, with mean hemoglobin A1c of 7.0 ± 0.9%.

**Table 1.** Baseline characteristics.

|  |  |
| --- | --- |
| *N* | 20 |
| Age (yr) | $48 \pm 18$ |
| Sex (Male/Female) | 11/9 |
| Race (White/Black) | 19/1 |
| BMI (kg/m<sup>2</sup>) | $29.5 \pm 5.2$ |
| Diabetes duration (yr) | $26 \pm 16$ |
| History of DKA (yes/no) | 7/13 |
| Hemoglobin A1c (%) | $7.0 \pm 0.9$ |
| Time in range 70-180 mg/dL (%) | $61 \pm 18$ |
| Insulin delivery (CSII/MDI) | 16/4 |
| Total daily insulin (units/kg) | $0.70 \pm 0.32$ |
| Estimated GFR (mL/min/1.73 m<sup>2</sup>) | 90 (83-94) |

BMI, body mass index; DKA, diabetic ketoacidosis; CSII, continuous subcutaneous insulin infusion; MDI, multiple daily injections; GFR, glomerular filtration rate. Data are expressed as mean ± SD or median (IQR) as appropriate.

The correlation between BrACE and BOHB during unsupervised outpatient care was moderate (ρ = 0.41; 95% CI: 0.36 to 0.45; *P* < 0.0001; **Figure 1**). During the DAPA period, the median (IQR) capillary blood BOHB concentration was greater than in the UC period [0.2 (0.1 – 0.3) mmol/L DAPA vs. 0.1 (0.1 – 0.2) mmol/L UC, *P* < 0.001]. Median (IQR) BrACE was also greater during the DAPA period than in the UC period [2 (1 – 4) ppm DAPA vs. 1 (1 – 3) ppm UC, *P* < 0.001]. During unsupervised outpatient UC periods, 3 of 718 BOHB readings were ≥ 1.5 mmol/L.

**Figure 1.**
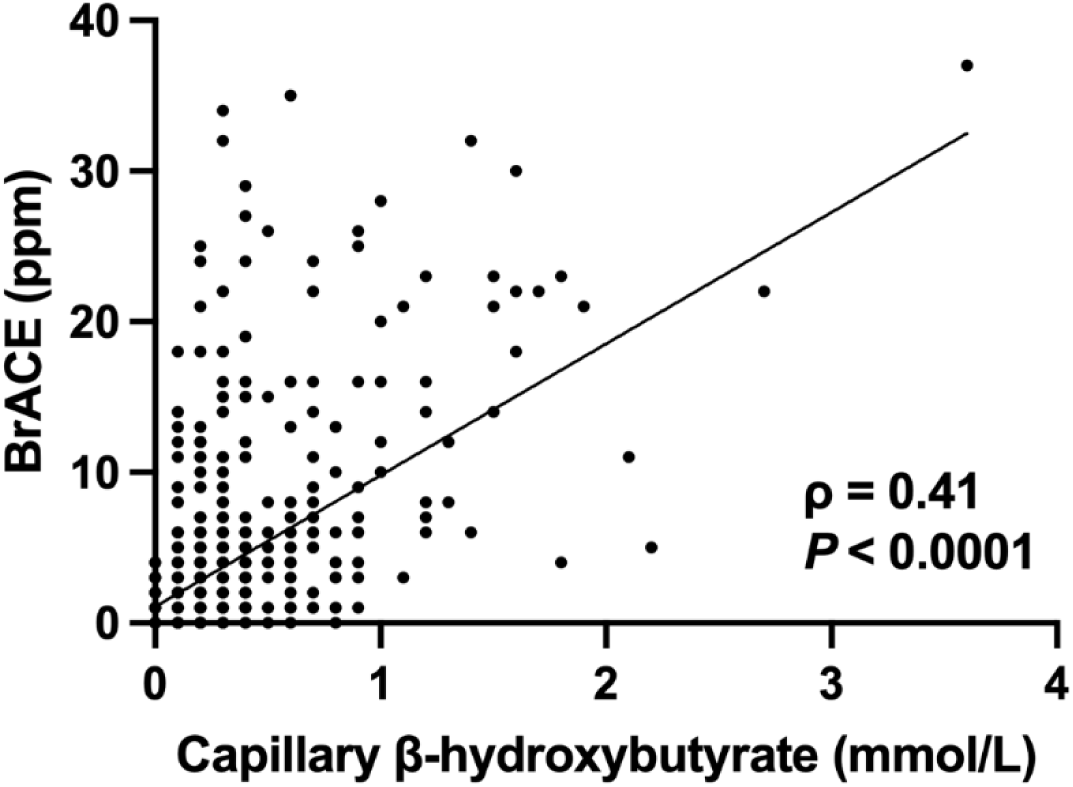
Correlation of blood and breath ketones during unsupervised outpatient periods. Paired capillary BOHB and BrACE measurements during unsupervised outpatient periods (n =1425 paired measurements). Linear trendline is shown.

During unsupervised outpatient DAPA periods, 10 of 707 BOHB readings were ≥1.5 mmol/L, but 6 of these occurred in a single bout of ketosis in one participant (**Figure 2**). In this individual, BrACE identified clinically significant ketosis during the outpatient dapagliflozin treatment period. Repeated paired BrACE and BOHB measurements showed that BrACE values were slower to normalize compared to capillary BOHB (**Figure 2**).

**Figure 2.**
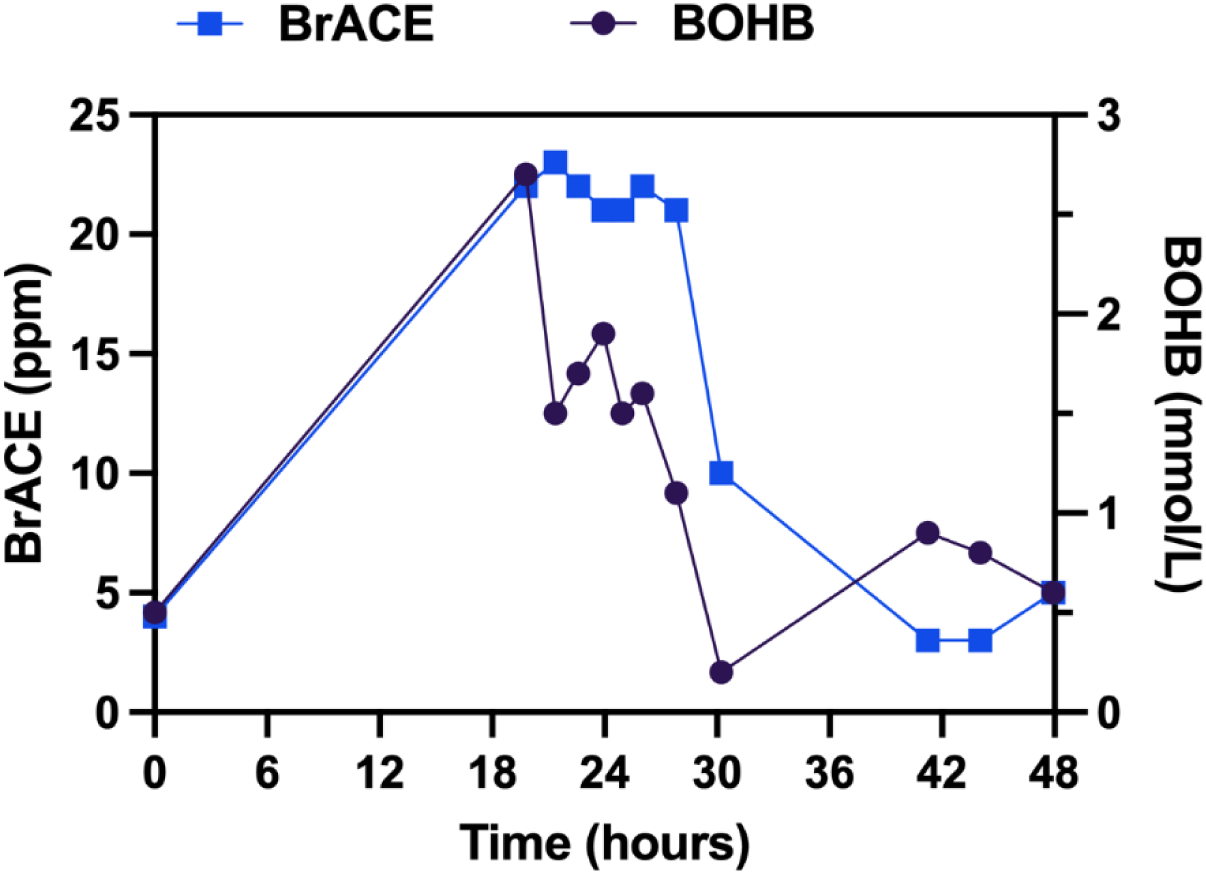
Paired BrACE and capillary blood BOHB values during a bout of outpatient ketosis. Paired values collected over a 48-hour period in a participant who developed and treated ketosis while taking dapagliflozin.

During the supervised insulin withdrawal visits, 341 paired capillary blood BOHB and BrACE measurements were collected. A strong correlation of capillary blood BOHB and BrACE during supervised insulin withdrawal was observed (ρ = 0.81; 95% CI: 0.77 to 0.84; *P* <0.0001; **Figure 3A**). The correlation of 254 paired plasma BOHB and BrACE measurements during supervised insulin withdrawal was similar (ρ = 0.82; 95% CI: 0.78 to 0.86; *P* <0.0001; **Figure 3B**).

**Figure 3.**
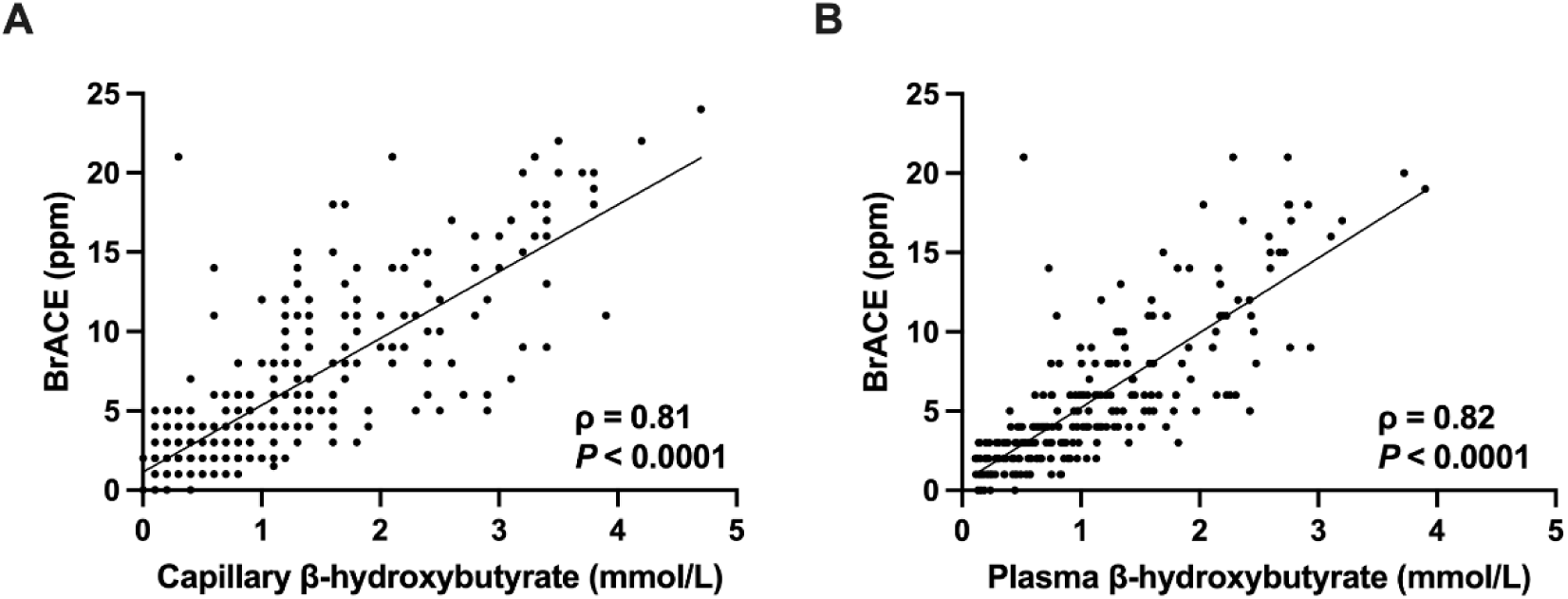
Paired BOHB and BrACE measurements during supervised insulin withdrawal. (A) Correlation of capillary blood BOHB and BrACE (n=341 paired measurements). (B) Correlation of plasma BOHB and BrACE (n=254 paired measurements). Study visits with and without dapagliflozin treatment were pooled (n=38 visits). Linear trendlines are shown.

When the data from the two-week unsupervised outpatient care and supervised insulin withdrawal study periods were combined, the correlation of capillary blood BOHB and BrACE was moderate (ρ = 0.56; 95% CI: 0.53 to 0.59; *P* <0.0001; **Figure 4**).

**Figure 4.**
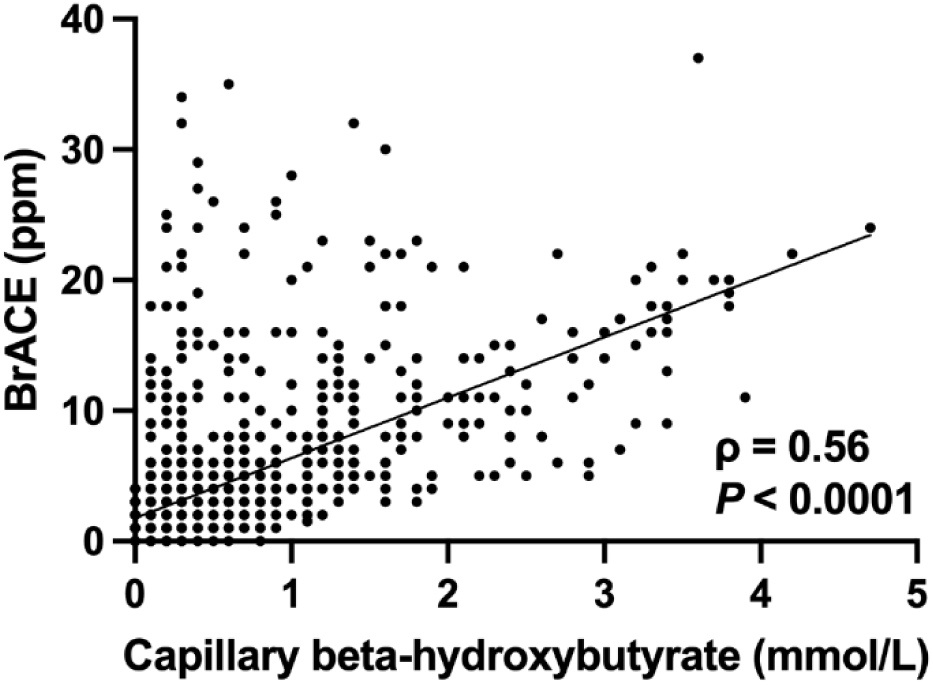
Correlation of capillary BOHB and BrACE during unsupervised outpatient care and supervised insulin withdrawal. Data from Fig. 1 and Fig. 3A were pooled (n=1766 paired measurements). Linear trendline is shown.

ROC analysis including data from both the unsupervised outpatient periods and the supervised insulin withdrawal visits with and without dapagliflozin was performed to determine the optimal BrACE value for identification of ketosis, defined as capillary BOHB ≥1.5 mmol/L (**Figure 5**). Optimal sensitivity (92%) and specificity (83%) were obtained at BrACE values ≥5 ppm.

**Figure 5.**
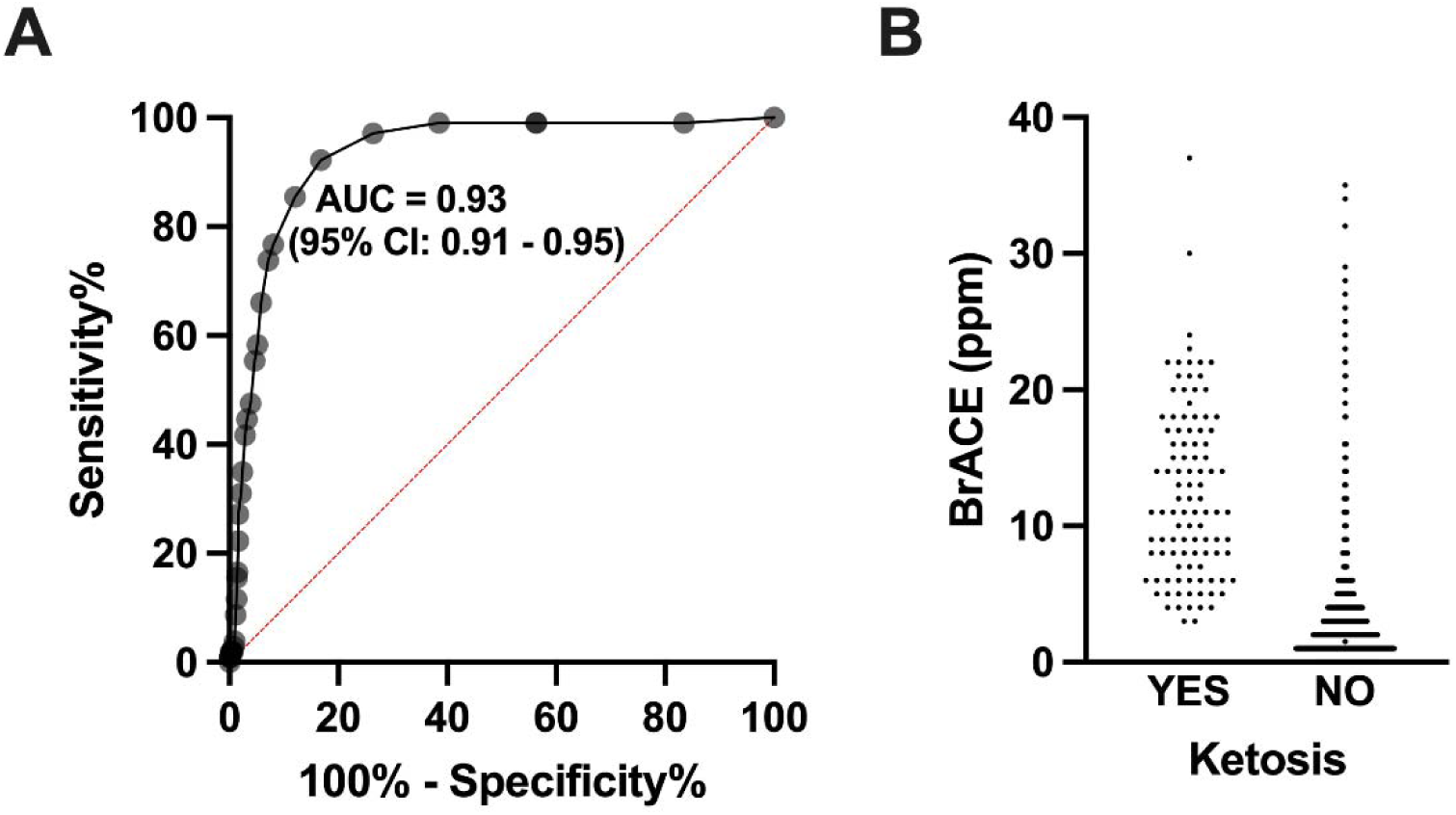
ROC curves for BrACE identification of ketosis. Pooled data from 2-week outpatient periods and supervised insulin withdrawal studies with and without dapagliflozin treatment were analyzed. Ketosis status was defined as capillary blood BOHB ≥1.5 mmol/L (YES) or <1.5 mmol/L (NO). n=1743 non-ketotic and 103 ketotic paired measurements.

As expected, dapagliflozin use in those with T1D was associated with a reduction in insulin requirements and an improvement in blood glucose metrics (**Table 2**). As compared to UC, DAPA was associated with a ∼11% reduction in total insulin requirement, a ∼5% decrease in average CGM glucose level and a ∼8% increase in time in range (70-180 mg/dL) **(Table 2**).

**Table 2.** Effect of dapagliflozin on insulin requirement and glycemia.

|  | UC (n=18-19) | DAPA (n=18-19) | P value |
| --- | --- | --- | --- |
| Daily basal insulin (units) | 27.2 (17.4 – 55.4) | 21.9 (15.4 – 40.9) | 0.0134 |
| Daily bolus insulin (units) | 31.4 ± 15.8 | 26.7 ± 11.6 | 0.0401 |
| Total daily insulin (units) | 47.9 (36.2 – 85.5) | 44.7 (35.0 – 76.3) | 0.0082 |
| Total daily insulin (units/kg) | 0.66 ± 0.30 | 0.59 ± 0.24 | 0.0104 |
| Average CGM glucose (mg/dL) | 154 (138 – 165) | 143 (139 – 147) | 0.0016 |
| Time-in-range 70-180 mg/dL (%) | 70.1 ± 10.7 | 75.6 ± 9.5 | 0.0111 |

Data are from each 14-day outpatient period, with or without dapagliflozin. CGM, continuous glucose monitor. UC, usual care. DAPA, usual care plus dapagliflozin. Data are mean ± SD or median (IQR) as appropriate. 18 of 20 subjects had paired insulin data and 19 had paired CGM data from both study periods for analysis. P values by Wilcoxon matched-pairs signed-rank test or paired two-tailed t-test.

## Discussion

The increased ketoacidosis risk associated with SGLT2i has prevented their regulatory approval and widespread use in people with T1D. To alleviate this concern and potentially allow those with T1D to benefit from the established cardiovascular and renal benefits of SGLT2i, expert consensus panels have recommended the development of noninvasive ketone monitoring technologies.^24^ Handheld BrACE analyzers represent one such technology with potential clinical utility. BrACE has been shown to correlate well with capillary blood ketone measurements in free-living people with T1D, but the performance of breath ketone analyzers has not been examined during insulin withdrawal—a scenario closely mimicking real-world ketogenesis associated with insulin pump failure or insulin omission.^32^

In this study, to explore the utility of BrACE measurement in T1D, participants monitored ketones with paired blood and breath assessments both during 14 days of their usual care at home, and during 14 days of usual care plus dapagliflozin treatment, which we hypothesized would enhance ketone production. Further, participants underwent supervised insulin withdrawal during usual care and immediately after 14 days of dapagliflozin treatment. We observed a strong correlation between capillary blood BOHB and BrACE during supervised insulin withdrawal conditions but a moderate correlation in the unsupervised outpatient settings.

The lower correlation between capillary blood BOHB and BrACE observed during the unsupervised outpatient periods may be related to several factors. First, most ketone values collected during the unsupervised outpatient periods were very low, generating the problem of restricted range.^36^ Second, proper use of the breath ketone analyzer including the correct breathing pattern and the use of a filter cap was enforced by supervision during the insulin withdrawal study. Third, VOC interference may have occurred in the outpatient setting. VOCs are released from many items used in daily life and can result in falsely elevated BrACE readings. During the insulin withdrawal study, all measurements were done during fasting and under supervision, so the technical validity of the measurements and the range of ketones produced were both greater.

In ROC analysis, a BrACE level of ≥5 ppm had 92% sensitivity and 83% specificity for detecting a capillary BOHB of ≥1.5 mmol/L. This level of BrACE was therefore considered the actionable level for prevention of ketoacidosis. A recent study of SGLT2i treatment in people with T1D demonstrated that DKA mitigation strategies including ketone monitoring with capillary BOHB or urinary ketones can reduce DKA episodes despite higher average BOHB levels in those taking sotagliflozin.^19^ BrACE measurement using a BKA is non-invasive, allows repeated measurements without additional cost and may represent a viable method for ketone monitoring that can be included in DKA mitigation strategies for persons with T1D using SGLT2i and those at increased risk of episodic DKA.

One individual experienced a clinically significant bout of ketosis during the unsupervised outpatient DAPA period. This individual did not eat breakfast and lunch, thus reducing his automated insulin delivery. He did not have hyperglycemia, did not manually change insulin doses, and did not report symptoms during the ketosis episode. He reported the rise in ketones to the study team and was instructed in a treatment strategy that led to resolution of the ketosis. He did not skip any further meals and did not have additional ketosis episodes during the remainder of the study period despite continuing SGLT2i treatment. Identifying persons at risk for episodic ketosis may require trial periods of SGLT2i use with frequent ketone monitoring and underscores the importance of appropriate patient selection for SGTL2i use in people with T1D.

Notable aspects of this study’s design include the collection of over 1700 paired BrACE and BOHB measurements and testing in the participants’ home environment. The data from the supervised insulin withdrawal study is relevant to scenarios such as insulin pump failure or loss of insulin access, in which absolute insulin deficiency gradually develops.^37^ This is important as CSII remains a risk factor for development of DKA.^38^

This study has important limitations including the small sample size. Further, in the unsupervised outpatient care periods, the range of values used for correlation calculations was relatively narrow and the risk for interference of VOC with BrACE measurements was higher. The restricted range likely contributed to the weaker correlation between BrACE and capillary BOHB observed in the outpatient setting. In addition, the strong correlation between BrACE and BOHB measurements during supervised insulin withdrawal may have been enhanced by the controlled environment of the research unit, in which paired measurements were supervised by a nurse and the BKA device was capped when not in use to reduce the risk of VOC interference. Additionally, this study has the limitation of excluding individuals with history of frequent DKA, uncontrolled diabetes (HbA1c >10%) or use of a ketogenic diet, who may be at increased risk for DKA with use of SGLT2i especially during insulin withdrawal. Future studies should consider investigating BrACE monitoring in patients at greater risk for ketoacidosis.

Our study demonstrates that BrACE measurement using a noninvasive, handheld breath ketone analyzer can detect clinically actionable ketosis but is more reliable in supervised settings as compared to unsupervised outpatient care. A critical limitation for more widespread real-world use in T1D is that currently available BKA devices remain highly susceptible to VOC interference. Whether repeated training on proper use of the device or deployment of a filter cap in the outpatient care setting would have reduced presumed interference in the BrACE measurements is unknown. Additional BKA device development is needed to explore modifications that could address these issues, making them more viable for home use in people with T1D. Additionally, the occurrence of ketosis in more than one patient while at home reminds health care providers that persons with T1D are at risk for asymptomatic ketosis that may lead to serious adverse consequences. The advice to test for ketones after symptoms develop may be too late for early detection and effective treatment at home.

## Conclusions

BrACE monitoring in adults with T1D offers a noninvasive assessment of circulating ketones. A BrACE reading of ≥5 ppm had 92% sensitivity and 83% specificity for a capillary blood BOHB concentration ≥1.5 mmol/L. The efficacy of the breath ketone analyzer is limited by common interfering VOC found in the home and by the need for proper operating technique. Larger studies are needed to further evaluate the utility of BrACE measurements as a substitute for capillary BOHB measurements in persons with T1D at risk of ketosis.

## Data Availability

All data produced in the present study are available upon reasonable request to the authors

## Abbreviations

(BrACE): Breath Acetone
(BOHB): β-hydroxybutyrate,
(T1D): Type 1 Diabetes,
(SGLT2i): Sodium Glucose Cotransporter-2 inhibitor,
(DKA): Diabetic Ketoacidosis,
(VOC): volatile organic compound,
(MDI): Multiple Daily Injections
(CSII): Continuous Subcutaneous Insulin Infusion
(HbA1c): Hemoglobin A1c
(eGFR): Estimated Glomerular Filtration Rate
(MOS): Metal Oxide Sensor
(CGM): Continuous Glucose Monitor
(BKA): Biosense Ketone Analyzer
(CTRU): Clinical Translational Research Unit

## Acknowledgments

We would like to thank the patients with T1D who participated in the study. We thank the staff of the Washington University Clinical Translational Research Unit and Core Lab for Clinical Studies for conducting the study visits and completing the sample analyses. We would also like to thank Readout Health for planning and technical assistance.

